# Precipitating factors and avoidability of 7-day readmissions after hospitalization for heart failure

**DOI:** 10.1101/2025.10.08.25337521

**Authors:** Cathy A. Eastwood, Nicholas VanKampen, Danielle Fox, Chelsea Doktorchik, Robin L. Walker, Hude Quan, Jonathan Howlett, Kathryn M. King-Shier

## Abstract

**Background:** Hospital readmissions within 7 days after discharge are considered highly avoidable and undesirable for the patient and hospital. It is important to clarify gaps in the quality of inpatient care to identify strategies to address this problem.

**Aims:** We aimed to describe the precipitating factors and determine the potential avoidability of 7-day readmissions after hospitalization for heart failure (HF).

**Methods:** A health record audit was undertaken of patients discharged after hospitalization for HF from Calgary, Alberta hospitals. Content analysis was undertaken to identify factors precipitating readmission, and readmission’s avoidability was qualitatively examined and scored based on descriptive categories.

**Results:** Of 18,590 patients admitted to the hospital for HF during the study period, 191 HF patients were readmitted within 7 days (50% female; mean age 78 years). Potentially avoidable readmissions (57%) were due to unresolved symptoms, unaddressed social or self-care issues, adverse events from the index admission, high disability without added services, and discussions of palliative care without added services. Readmissions deemed less avoidable (43%) were due to new health issues, recurring symptoms post-stability at discharge, or refusal of care.

**Conclusion:** Only half of hospital readmissions within 7 days after HF discharge were related to HF and more than half were scored as avoidable. We provide novel criteria for identifying the avoidability of 7-day readmissions that could be used for assessing HF patients’ readiness for discharge and potentially reducing readmission rates.

## Background

Hospital readmission stands as a crucial indicator of disease severity and healthcare quality globally, especially for prevalent chronic conditions like heart failure (HF). With over 64 million individuals worldwide affected by HF, it poses a significant health challenge, contributing to more than 1 million hospitalizations in the United States and Europe alone (1–3). The global economic burden of HF, estimated at approximately U.S. $346 billion annually, underscores the substantial impact of HF-related hospitalizations (2). Despite advancements in treatment, HF remains a prominent cause of hospital readmissions. Factors such as nonadherence to treatment plans, lack of education, and limited access to care contribute to poorer outcomes. A recent systematic review (4) covering >1.4 million people with HF across 18 countries found a pooled average 30-day readmission rate of 13% (95% CI: 10%-16%) worldwide. In Canada specifically, an estimated 20% of patients experience readmission within 30 days of discharge (5). Notably, a significant portion (6-10%) of these readmissions occurs within 7 to 14 days post-discharge (6). Readmissions within 7 days of discharge are more actionable and more related to what occurred during the hospitalization. Given the potential avoidability of 7-day readmissions, there is a growing imperative to investigate the factors contributing to such occurrences (7, 8).

A good portion of readmissions in the HF population result from the natural worsening of the disease or events outside the control of healthcare (9). However, there is a growing consensus among researchers that distinguishing and evaluating avoidable readmissions could offer a more meaningful metric for assessing the quality of care (10). While previous studies have primarily focused on determining the avoidability of readmissions within general medical-surgical populations using subjective criteria (11, 12), there remains a lack of research focusing on the one-week post-discharge period and the assessment of avoidability in HF populations (13). Therefore, understanding the factors contributing to readmissions, particularly within the HF population, is essential for developing targeted quality improvement strategies.

The objective of this study was to investigate 7-day readmissions following hospitalization for HF, aiming to delineate 1) the presenting problems upon readmission, 2) the contributing factors to readmission, and 3) the type of readmissions that might be avoidable. By clarifying these aspects, we seek to provide insights that can inform the development of effective interventions and enhance the quality of care provided to HF patients.

## Methods

From a comprehensive health record audit of 7-day all-cause readmissions after HF hospitalization, we conducted a detailed qualitative analysis of clinical, health system, and patient-related events leading to readmission (3,10). The Conjoint Health Research Ethics Board of Alberta approved this study. The investigation conforms with the principles outlined in the Declaration of Helsinki (14).

### Study Cohort

Of 18,590 patients admitted for HF in Alberta between 2004-2012 in an earlier study (10), 234 were readmitted within 7 days of discharge to a hospital in Calgary, AB. These 234 charts were scrutinized and 191 were confirmed as non-elective readmissions, which became the study cohort. In brief, all patients were hospitalized at three acute care hospitals in Calgary, Alberta, Canada with a primary diagnosis of HF (International Classification of Diseases, Tenth Revision (ICD-10-CA) code I50.x) between 1 April 2004 to 31 March 2012. Inpatient and outpatient cardiovascular and HF services are available at these hospitals. Cases were excluded if not an Alberta resident, age <19 or >105 years, in-hospital death (or had died following the index admission, and were not readmitted), or discharged to another acute care facility. If repeated discharges from the hospital occurred and were followed by readmission within 7 days, the records were included as separate cases to capture the maximum number of readmissions occurring within 7 days of discharge.

### Data Collection

From the index admission health records, the investigator and two research assistants (CE and DF, both nurses) extracted demographic data about each patient, detailed descriptions of their health status at discharge, and events leading up to readmission. Readmission record data included symptoms and chief complaints from the emergency department notes, history and physical findings, primary diagnoses, treatment course, and disposition from the discharge summary. Along with the presenting symptoms, the researchers noted documented actions taken by the patient or family in response to the symptoms and the actions taken by healthcare providers in response to the symptoms before readmission and upon hospitalization. Narrative summaries were written on the data collection forms in as much detail as possible, which became the qualitative text for analysis.

### Data Analysis

Data were analyzed by the investigator and one research assistant (cardiology staff nurse) using conventional content analysis (15). The intention was to cluster and summarize the main reasons for readmission, risk factors, health system processes, and events in plain language with minimal interpretation (16). Each team member made notes identifying initial thoughts and prominent keywords. These data were grouped into categories and were given coded labels.

To ensure agreement on each larger category and subgroup, the researchers jointly reviewed all 191 readmission cases and reached a consensus when discrepancies were present. After reaching a consensus, codes were clustered into larger categories, which included: 1) the readmission presenting health problem, 2) contributing factors, and 3) the degree of avoidability. Subcategories were noted under these larger headings. The qualitative text and subcategory groups were read again by both team members to ensure the most prominent factor was identified. Only one subcategory label was assigned for each readmission case if more than one condition was present (e.g., urinary infection and heart failure). Labels emerged inductively during coding.

### Scoring Avoidability

Avoidability was scored using 2 approaches, implicit and explicit review. *Implicit* review (Table 1) and *explicit* review (Table 2) were undertaken by 3 team members.

**Table 1:**
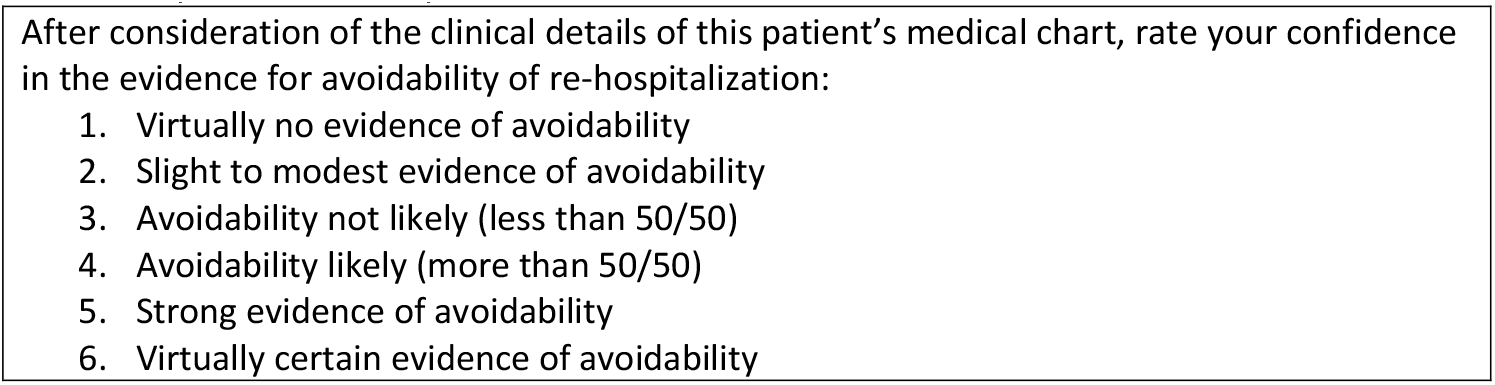
Implicit avoidability criteria.

**Table 2:** Explicit avoidability criteria.

|  |
| --- |
| **Low Avoidability 1 to 3** |
| • New, unforeseen problem not related to HF or a condition present during index admission<br>• Refusal of care; left against medical advice during index admission<br>• Same signs and symptoms as during index admission but was stable at discharge<br>• Services provided but not enough (e.g., homecare was arranged but patient went to hospice or long-term care) |
| **High Avoidability 4 to 6** |
| • Presence of the same signs and symptoms not resolved at index discharge<br>• Discussion of palliation but no added palliative services, hospice, or change in goal of care<br>• High disability but no extra home services or increased level of disposition to long-term care, etc.<br>• Social or self-care issue not addressed with extra services<br>• Adverse event related to clinical care during index admission |

During data collection, the avoidability of readmissions was evaluated by implicit review of the health record using a 6-point scale (Table 1). Implicit review involved using professional judgment to determine a score for avoidability on a 6-point scale with minimal predetermined criteria. This method has been used to evaluate the causation and preventability of adverse events and readmissions after hospitalization for general medical and surgical diagnoses(11, 17). Iterations of the scoring avoidability tool were assessed on inter-rater reliability among 3 researchers. We included minimal wording for each criterion on the 6-point scale to reduce variation. For example, low avoidability (1-3 on the scale) would be assigned if a new problem occurred for the patient; high avoidability (4-6 on the scale) would be assigned if the patient returned for the same problem.

Explicit review involved developing novel criteria for a 6-point rating scale after examining the data, and the literature, and reaching agreement among team members. Explicit criteria for low avoidability and high avoidability of readmissions were developed based on past literature (18–21). Subjective wording (e.g., premature discharge; inadequate palliative care) was minimized and more description was added for each level based on patterns revealed while reviewing with implicit criteria. The investigator and research assistants scored avoidability again using the explicit criteria (Table 2). Avoidability was scored a 3 if some criteria for low avoidability were present but avoidability was deemed not likely (<50/50). Avoidability was scored 4 if some criteria for high avoidability were present and avoidability was deemed likely but not certain (>50/50).

## Results

Of 18,590 patients admitted to the hospital for HF during the study period, 191 HF patients were readmitted within 7 days (50% female; mean age 78 years). Forty-eight (26%) lived alone while the others lived with a spouse, family member, or in a care facility. Details on comorbid conditions and clinical characteristics have been previously published (10).

### Presenting Pathology at the Time of Readmission

A wide variety of factors contributed to readmission within 7 days of discharge. The pathologic conditions that brought patients back to the hospital varied greatly, though clusters appeared. Most patients presented with symptoms other than HF (69%) (Figure 1). Other cardiac conditions (23%) such as atrial fibrillation, bradycardia, and chest pain were common. Gastrointestinal (GI) disorders (13%) included diarrhea, GI bleeding, ischemic bowel, and other GI disorders such as undiagnosed abdominal pain. Respiratory issues (13%) included pneumonia, upper respiratory infections, or chronic obstructive pulmonary disease (COPD). Urinary and renal issues (5%) were predominantly infections. Other conditions (18%) included altered mental status, mobility issues, electrolyte or metabolic imbalances, other infections (e.g., cellulitis), cancer, stroke, coagulation issues, and liver disease (e.g., hepatic encephalopathy). Two patients presented with systemic reactions to oral medications.

**Figure 1:**
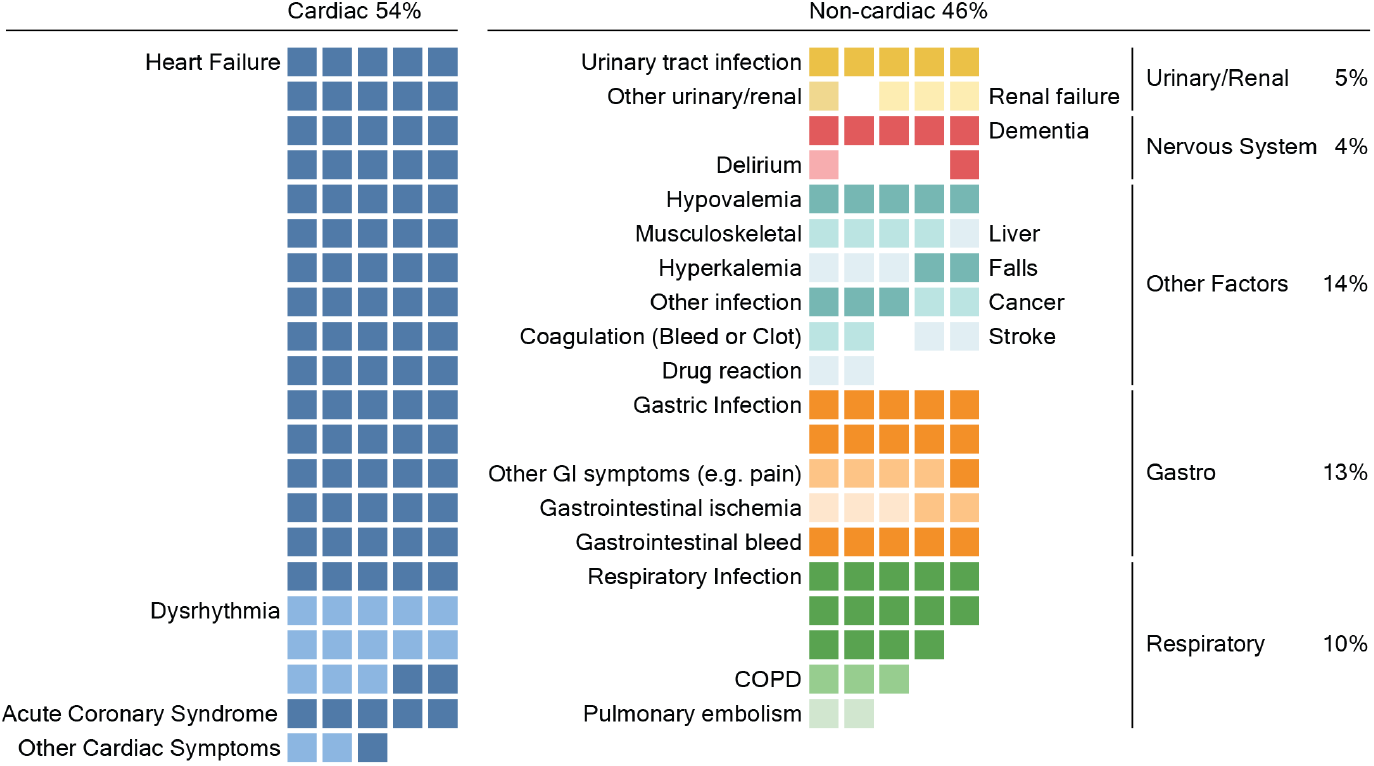
Presenting health problem at the time of readmission within 7 days of discharge. *Note:* One box represents one patient. GI = gastrointestinal, COPD = chronic obstructive pulmonary disease

### Factors Contributing to the Readmission

When documented, we identified the process or event that may have triggered the symptoms and related pathology that required readmission (Table 3). Sixty percent (115/191) of the readmission records had symptoms documented but minimal or no description of what may have precipitated the symptoms. Of this subgroup, a large portion of readmissions was due to continuing symptoms from the index admission. Continuing symptom descriptions include: “short of breath since discharge”, “throbbing headache since discharge”, “exacerbation related to recent MI and resulting pericarditis”, or “readmitted for abdominal pain, continuous since discharge yesterday”. New health conditions made up the next largest category including gastrointestinal bleeds, bradycardia, confusion, or falls. Of these two subgroups, there was not enough information to discern what may have contributed to the development of the symptoms.

**Table 3:** Principal factors contributing to readmission.

| Factors (n=191) | Description and examples |
| --- | --- |
| Continuing symptoms not resolved at discharge n=80 (42%) | HF symptoms (e.g., dyspnea, edema)<br>Other continuing symptoms also present during index admission (e.g., dysrhythmia, epistaxis, COPD, pneumonia, pulmonary fibrosis, knee pain, diarrhea) |
| New health conditions n=35 (18%) | New symptoms, no link to prior admission (e.g., “came to ED with abdominal pain and nausea”, chest pain, “found confused by daughter, had fallen twice, bradycardia, low glucose”, cardiac arrest, hyperkalemia) |
| Medication-related n=20 (10%) | Adverse drug reaction (e.g., hives from antibiotic)<br>Over-medicated (e.g., digoxin toxicity, fell out of bed after taking sleeping pill)<br>Under-medicated (e.g., no HF meds prescribed) |
| Self-care issue n= 16 (8%) | Patient non-adherence to prescribed regimen (e.g., CPAP not worn, prescriptions not filled by family, patient stopped self-care |
|  | due to mood disorder, patient stopped medications due to feeling well)<br>Misunderstanding the care plan (e.g., excessive fluid intake, ate high sodium soups, used old medication list)<br>Self-harm (e.g., overdose of 10 Tylenol #4 tablets) |
| Refusal of care n=10 (5%) | Left against medical advice<br>Declined homecare or long-term care<br>Refused functional or cognitive assessment |
| Potentially inappropriate placement n= 9 (5%) | Mobility issues (e.g., unable to get off toilet)<br>Increased confusion/dementia (e.g., combative at home)<br>Current services not enough (e.g., “difficulty coping despite current maximum homecare”); spouses couldn’t manage patient at home anymore (e.g., patients had metastatic cancer, somnolence, dyspnea, and anxiety too great to cope at home) |
| Failure to thrive n= 11 (6%) | Weakness, gradual functional decline (e.g., dehydration from poor oral intake, not eating or drinking for days, high risk for falls with unsteady gait, severe lethargy and/or weakness, progressive dementia)<br>Decline in status despite added services |
| Hospital-related n=5 (3%) | Complication of recent procedure or therapy (e.g., arm cellulitis secondary to intravenous trauma, nosocomial pneumonia, clostridium difficile colitis, pulmonary embolus post pacemaker insertion) |
| Palliative n= 4 (2%) | Active dying (e.g., unable to eat/drink and died in hospice 4 days post readmission, advanced age with severe multisystem symptoms, advanced cancer, died during readmission) |
| Healthcare provider error n=1 (<1%) | Poor standard of community care (e.g., “nursing home let oxygen tank run out and patient became hypoxic”) |

COPD = chronic obstructive pulmonary disease; ED = emergency department; CPAP =
continuous positive airway pressures

Of presenting patient symptoms upon readmission, 40% (76/191) of the cases could be linked to health system processes or patient-related factors. Health system processes appeared to contribute to 21% of the readmissions. These cases involved challenges with medications (e.g., digoxin toxicity or no HF medications prescribed), inappropriate placement (e.g., discharged home with severe immobility or dementia), hospital-related complications (e.g., “cellulitis related to intravenous”), or healthcare provider error (e.g., “oxygen tank ran out in long-term care facility”). In cases where palliative care was the focus, hospice or home-based palliative services may have been more suitable than an acute care readmission; thus, the readmission was categorized as a health system factor and not a patient-related factor.

Patient-related factors (19%) included self-care issues such as non-adherence to the prescribed regimen, misunderstanding the care plan, and self-harm. Patient factors also included the refusal of care or a swift decline in function in elderly patients described as “failure to thrive”.

### Avoidability

When avoidability was ranked using implicit criteria, the greatest number of cases (75% 143/191) were scored in the lower avoidability categories (score 1-3) (Table 4).

**Table 4:** Potential avoidability of readmission scored using implicit criteria.

| Avoidability | Implicit n (%) |
| --- | --- |
| 1. Virtually no evidence of avoidability | 33 (17.3) |
| 2. Slight to modest evidence of avoidability | 71 (37.2) |
| 3. Avoidability not likely (less than 50/50) | 39 (20.4) |
| 4. Avoidability likely (more than 50/50) | 33 (17.3) |
| 5. Strong evidence of avoidability | 15 (7.9) |
| 6. Virtually certain evidence of avoidability | 0 |
| Total | 191 (100) |

When avoidability was ranked on a 6-point scale using explicit criteria, the largest cluster (56%, 107/191) scored with strong evidence of avoidability (score 4-6) (Table 5). This designation was assigned to patients who returned with the same unresolved symptoms present during the index admission, were not provided with more post-discharge services when demonstrating evidence of high disability, had social issues that were not addressed with added services, or encountered an adverse event related to clinical care during the index admission.

**Table 5:** Description of reasons for potential low and high avoidability (explicit criteria)

|  |  |
| --- | --- |
| *Low Avoidability (n/%)* |  |
| • New unforeseen problem | 39 (21%) |
| • Continuing symptoms but stable at discharge | 32 (17%) |
| • Refusal of care | 10 (5%) |
| • Services provided but not enough | 3 (2%) |
| Subtotal | 84 (44%) |
| *High Avoidability* |  |
| • Signs and symptoms present at discharge | 42 (22%) |
| • Adverse event related to clinical care | 23 (12%) |
| • Social or self-care issue not addressed with added services | 20 (11%) |
| • High disability but no added services | 16 (8%) |
| • Discussion of palliation but no added services | 6 (3%) |
| Subtotal | 107 (56%) |

The other 44% (84/191) cases were deemed less avoidable (scored 1-3) due to a new unforeseen problem, a recurrence of symptoms following being stable at discharge, refusal of care, or additional problems despite the provision of services. Examples of new unforeseen problems included cardiac arrest, chest pain, delirium, falls, or stroke. Many cases (40.6%, n/N) within these larger categories had unclear evidence of high or low avoidability and were ranked as 3 or 4.

### Deaths During Readmission

Deaths occurred in 14.7% (28/191) of patients during readmission. Of those, 4 patients were described as terminally ill or requiring palliative care in the readmission history and physical notes. Other patients who died during the readmission were classified as “failure to thrive”, exhibited continuing symptoms from the index admission, or had new unforeseen problems (e.g., stroke).

## Discussion

This study revealed that patient readmissions within 7 days of discharge were attributed to various causes, with 41% due to HF, 13% to other cardiac conditions, and 46% to non-cardiac reasons, including gastrointestinal, respiratory, urinary/renal, other physiological conditions Factors contributing to readmission unresolved symptoms, new health conditions, medication-related ailments, self-care challenges, refusal of care, inappropriate placement, failure to thrive, hospital-related complications, palliative care discussions, and health-care provider errors. Notably, over half (56%) of readmissions were considered potentially avoidable based on explicit criteria, while implicit review produced a more conservative estimate. Avoidable readmissions included patients with unresolved symptoms, unaddressed social or self-care issues, adverse events related to clinical care during index admission, and high disability or terminal illness without added services.

Two different mechanisms were used to determine the avoidability of readmissions that resulted in two different sets of scores. Other studies also found that evaluation of the quality of medical care was highly dependent upon the method used for measurement – implicit versus explicit review criteria (22). They described that when implicit criteria were used, the acceptability of medical care was judged more leniently and was more open to variation between judges than when using explicit criteria. Likewise in our study, when implicit criteria (professional opinion) were used, 25% of readmissions were scored as moderately to highly avoidable, while the detailed explicit criteria resulted in 74.8% scored as moderately to highly avoidable. More specific explicit criteria guided the reviewers to a more detailed review and discrimination between avoidable and non-avoidable scoring options.

Although physician panels have used implicit criteria to determine readmission avoidability(11), it was not sufficient for our team (3 nurses) to consistently judge without more explicit criteria. Goldfield (2011) suggested clinicians within institutions, not just researchers with access to physician review panels, must be able to use reproducible methods to evaluate the avoidability of readmissions to activate quality improvement initiatives(23). Researchers found explicit criteria useful to reduce the risk of misclassifying the avoidability of each readmission case (24). Further, van Walraven et al. (2011) found that the criteria for evaluating the avoidability of readmissions have been non-specific, resulting in great variation between reviewers’ opinions of avoidability (11). If HF readmission rates are to be linked with hospital performance indicators or clinicians’ ability to assess for risk of readmission during discharge planning, then our proposed explicit criteria may produce useful estimates of avoidable readmissions and warrant further testing.

There has been a great deal of attention to the importance of identifying, acting on, and being penalized for avoidable readmissions in the HF population. However, a real-time prospective approach to prevention of avoidable readmissions is still needed. Ashton et al. (1994) developed explicit criteria for discharge readiness of HF patients that have not been further tested (25). Our avoidability criteria, coupled with identification of frailty, could inform the development of a screening tool for discharge readiness for clinicians and patients.

A reduction in readmissions may be attainable with the right precautions, such as screening for unresolved issues or referral to other specialties that can resolve unaddressed issues before discharge. An earlier study demonstrated that the likelihood of a 7-day readmission following HF was attributed to two factors: 1) patient frailty, and 2) patients under the care of an inpatient specialist physician (26). There was a decreased likelihood of readmission if follow-up care with a family physician was documented within 1 week of original hospital discharge. Thus, it is reasonable to implement procedures such as screening for frailty and scheduling patients for immediate follow-up after discharge from the hospital would reduce the frequency of readmissions in the HF population.

Certain social factors play a role in readmission rates as well. For example, women with limited health literacy are more vulnerable to readmission than men with limited health literacy (27). Physician characteristics (e.g., patient volume) are also reported as associated with hospital readmission rates in that a physician’s hospital activity, as reflected by the number of patient discharges per year (>100) results in lower 7-day readmission rates (28). Further, HF patients who were discharged with HF services had decreased odds (OR= 0.67, CI= 0.54-0.82) of being readmitted for any cause in 7 days compared to those without HF services, suggesting that prevention of readmission is possible with HF services (29). Similarly, from the current study, ensuring patients are referred to other healthcare professionals for unmet needs at discharge (e.g., those with unresolved symptoms or severe disabilities) may reduce readmissions.

A notable strength of this study is the rigor of the content analysis methods used. We reported symptoms, events, and quotes with minimal interpretation. For accuracy, we trained team members on guidelines-based treatment of HF and each person spent 30 to 60 minutes per health record gathering details on what happened at the time of readmission. We enhanced dependability by independently reviewing and coding the narrative text before the results were compared and discussed. As well, we let time pass before reviewing the data and codes again to gauge stability over time. Codes and categories were compared with published studies.

Our study has limitations. Regarding generalizability, our findings are most applicable to other large urban centers as our data was only captured from Calgary, Alberta hospitals. However, given the prevalence of the problem of readmission internationally, readers may find our results highly relevant. We aimed to enhance generalizability by using a sample that included almost all HF patients readmitted within 7 days of discharge during the years of interest based on minimal exclusion criteria. Further, the quality of our results relies largely on the quality of hospital chart documentation. Given some variation in documentation quality, which is expected with most health records, our findings were fully described as possible. We were unable to include patients in the study who died outside of the hospital during the 7-day follow-up period after the index admission. We did, however, assess factors contributing to readmission in the sample that had died during readmission (n=28). Finally, our avoidability scale was not externally validated. We encourage future research to validate this scale for use in further studies.

## Conclusions

We identified factors that contribute to 7-day readmission and whether these factors were avoidable. Our team developed novel explicit criteria to score readmission avoidability. We found explicit criteria to be more sensitive than using implicit criteria. These findings have implications for improving patient care and ultimately reducing avoidable readmissions among HF patients. Future research should focus on validating the explicit avoidability scale to improve discharge planning (e.g. readmission risk screening tools).

## Data Availability

Data cannot be shared publicly because to respect the privacy of the participants. Data are available from the University of Calgary Institutional Data Access / Ethics Committee (contact via) for researchers who meet the criteria for access to confidential data.

## Acknowledgements

We acknowledge Dr. Søren Knudson for assisting with the graphic display of the results.

## Data Availability Statement

Data available upon request.

